# Use of antimicrobials during the COVID-19 pandemic: a qualitative study among stakeholders in Nepal

**DOI:** 10.1101/2023.06.23.23291835

**Authors:** Binod Dhungel, Upendra Thapa Shrestha, Sanjib Adhikari, Nabaraj Adhikari, Alisha Bhattarai, Sunil Pokharel, Abhilasha Karkey, Direk Limmathurotsakul, Prakash Ghimire, Komal Raj Rijal, Phaik Yeong Cheah, Christopher Pell, Bipin Adhikari

**Affiliations:** Central Department of Microbiology, Tribhuvan University, Kathmandu, Nepal; Manmohan Cardiothoracic Vascular and Transplant Center, Institute of Medicine, Tribhuvan University, Kathmandu, Nepal; Center for Tropical Medicine and Global Health, Nuffield Department of Medicine, University of Oxford, Oxford, United Kingdom; Oxford University Clinical Research Unit, Patan Academy of Health Sciences, Lalitpur, Nepal; Mahidol-Oxford Tropical Medical Research Unit, Faculty of tropical Medicine, Mahidol University, Bangkok, Thailand; Amsterdam Institute for Social Science Research, University of Amsterdam, Amsterdam, The Netherlands

**Keywords:** COVID-19 disease, COVID-19 pandemic, antimicrobial resistance (AMR), Over-the-counter (OTC), Self-medication

## Abstract

**Introduction:** The COVID-19 pandemic was a major public health threat and posed tremendous pressure to develop a cure for it. Apart from ongoing efforts in developing vaccines, a lot of empirical treatments were recommended, that may have expedited the use of antimicrobials. The main objective of this study was to explore if and how the pandemic posed pressure on antimicrobials in Nepal using semi-structured interviews (SSIs) among patients, clinicians and drug dispensers.

**Methods:** A total of 30 stakeholders (10 each among clinicians, dispensers and COVID-19 patients) were identified purposively and were approached for SSIs. Clinicians and dispensers working in three tertiary hospitals in Kathmandu were first approached and were asked for their support to reach out to COVID-19 patients who were on follow-up at their out-patient department. SSIs were audio recorded, translated and transcribed into English, and were analyzed for thematic synthesis.

**Results:** Over-the-counter (OTC) uses of antibiotics were widespread during the pandemic, and were mostly rooted to patients’ attempts to halt the potential severity due to respiratory like illnesses, and the fear of being identified as a COVID-19 patients. Being identified as a COVID-19 patient was feared because of the stigmatization and social isolation. Patients who visited the drug shops and physicians were reported to make demands on specific medicines including antibiotics that may have added pressure among physicians and dispensers. Clinicians reported a degree of uncertainty related to treatment and that may have added pressure to prescribe antimicrobials. All stakeholders, although mostly patients and dispensers with limited understanding of what constitutes antimicrobials and the mechanisms underpinning it reported that the pressure during the pandemic may have added to the adversities such as antimicrobials resistance.

**Conclusions:** COVID-19 added a pressure to prescribe, dispense and overuse antimicrobials and may have accentuated the pre-existing OTC use of antimicrobials. Future pandemics including infectious disease outbreaks are major public health incidents that warrant a special caution on inappropriate pressure on antimicrobials. Strict policies related to the use of antimicrobials are urgent to redress their use during normal and pandemic situations.

## Introduction

The viral pandemic caused by Severe Acute Respiratory Syndrome Corona Virus 2 (SARS-CoV-2) has affected 765 million people, resulting in 6.9 million deaths worldwide as of 5th May 2023 [1]. Because of the efforts made by scientific communities across the globe, effective therapies and vaccines were developed at an unprecedented speed [2, 3]. Nonetheless, diverse variants of the virus are emerging and constantly posing challenges, thereby resulting in decreasing but sustained morbidity and mortality [4].

Concurrently, the world faces the threat of antimicrobial resistance (AMR), which may have even more catastrophic health impacts in the long term. Although its impacts are less obvious now, drug-resistant pathogens are now responsible for approximately 700,000 deaths per year, with this potentially rising to 10 million deaths annually by 2050 [5, 6]. Although a naturally occurring process, the emergence and spread of antimicrobial resistance is accelerated by the use of antimicrobials. Obtaining antimicrobial over-the-counter (OTC) and self-medication hence contribute to this process, yet potentially offer little therapeutic benefit, particularly in the absence of appropriate diagnostics [7-9].

In the early stages of the pandemic, diverse therapies were used in response to the rapidly rising mortality [10] and several studies have highlighted the extensive use of antimicrobials [11]. Viral in nature, COVID-19 infections are untreatable by antibiotics, but complications include pneumonia, chronic obstructive pulmonary disease (COPD), mucormycosis and other nosocomial infections, which are driven by bacterial and fungal pathogens and require case management with antibiotics and/or antifungals [10].

Globally and in Nepal health systems had enormous impact and that inevitably affected the health service delivery [12, 13]. With strict lockdown and disrupted healthcare systems, OTC acquisition of drugs and self-medication is thought to have increased during the pandemic [4, 14]. People with perceived COVID-19 and/or those who feared infection resorted to various measures and substances, including traditional and OTC medicines [15]. This was particularly pronounced in community settings in low- and middle-income countries (LMICs), where antimicrobials are often used without a laboratory diagnosis with an antimicrobial susceptibility assay [16-19]. Moreover, LMICs are especially vulnerable to the added burdens of AMR, owing to the constraints in resources, poor disaster preparedness health care services, poor governance, and lack of effective regulatory and legislative bodies [20].

With efforts and resources diverted towards the control and management of COVID-19 infections, the pandemic’s potential impact on the use of antimicrobials and potential rates of AMR has often been overlooked [21]. Given its potentially critical impact, few studies have sought to understand how the COVID-19 pandemic affected the use of antimicrobials. Fewer have used qualitative research methods to examine the underlying social, cultural and health system contributors of antimicrobial use during this period. Drawing on SSIs with diverse stakeholders, including patients, clinicians and drug dispensers, this article examines the use of antimicrobials, during the pandemic in Kathmandu, Nepal.

## Materials and Methods

### Study Design

An exploratory cross-sectional study was conducted among three broad stakeholders. The study drew on in-depth SSIs with clinicians who have treated COVID-19 patients, drug dispensers and COVID-19 patients. Patients who were confirmed to have been diagnosed with COVID-19 based on the national COVID management guidelines were considered as respondents in this study. COVID-19 guidelines confirmed the diagnosis based on the polymerase chain reaction (PCR) at test allocated centers. Reflecting on their experience with the COVID-19 event, the study followed broadly a phenomenological theoretical approach. The study followed a COREQ (Consolidated criteria for Reporting Qualitative research) guideline (**Supplementary File 1**).

### Study Settings

This study was conducted in and around three major tertiary hospitals in Kathmandu, Nepal. Kathmandu is also most populated urban area and experienced the highest COVID-19 case load and fatalities during the pandemic [22]. The three hospitals were purposively selected for this study that included Sukraraj Tropical and Infectious Disease Hospital (only hospital in the country that is dedicated for infectious diseases), Bir Hospital (the oldest tertiary hospital in the country), and Norvic International Hospital.

Around these hospitals, there were a variety of private healthcare care providers, including dispensaries, which were small privately-owned pharmacies with some offering clinical services and were responsible for wide coverage of healthcare services. While some dispensaries arranged appointments for patients with clinicians, the majority functioned as drug outlets.

### Sampling procedure, recruitment and semi-structured interviews

This study employed the purposive sampling method to select respondents. Participants were selected from the chosen hospitals for this study. At first, a clinician from each of the hospitals was approached and were asked for their suggestions for the patients. Similarly, dispensaries around the hospital and community pharmacies away from the tertiary facilities were approached for the diversity of respondents. Clinicians were recruited based on their exposure to and interactions with COVID-19 patients. Pharmacists or drug dispensers (also known as drug sellers or salespersons) who had qualifications to sell drugs at the pharmacy were selected in the study. Potential COVID-19 patients were recruited based on the history of being COVID-19 positive and seeking treatment at any point during the pandemic. All respondents were acquainted during the process of the study. The first author (BD) had access to information of respondents that could help identify individual participants for an interview.

A total of 30 respondents participated in this study that included 10 each from clinicians, drug dispensers and patients and took place between October 2022 and February 2023 (**Table 1**). Five dispensers among the approached population refused to take part in the study, only two of them expressed their reasons for refusal to participate, and the main reason was that they suspected the interviewer could be an inspection authority or a journalist. Two clinicians could not participate as promised by them because of their busy schedules and one patient was taking part in SSIs but had to leave for his home during the middle of the conversation. The number of respondents per each category was deemed adequate based on the content of the interviews where no new information was found after additional interviews [23]. Further informal discussions were held among the additional respondents who echoed the findings.

**Table 1:**
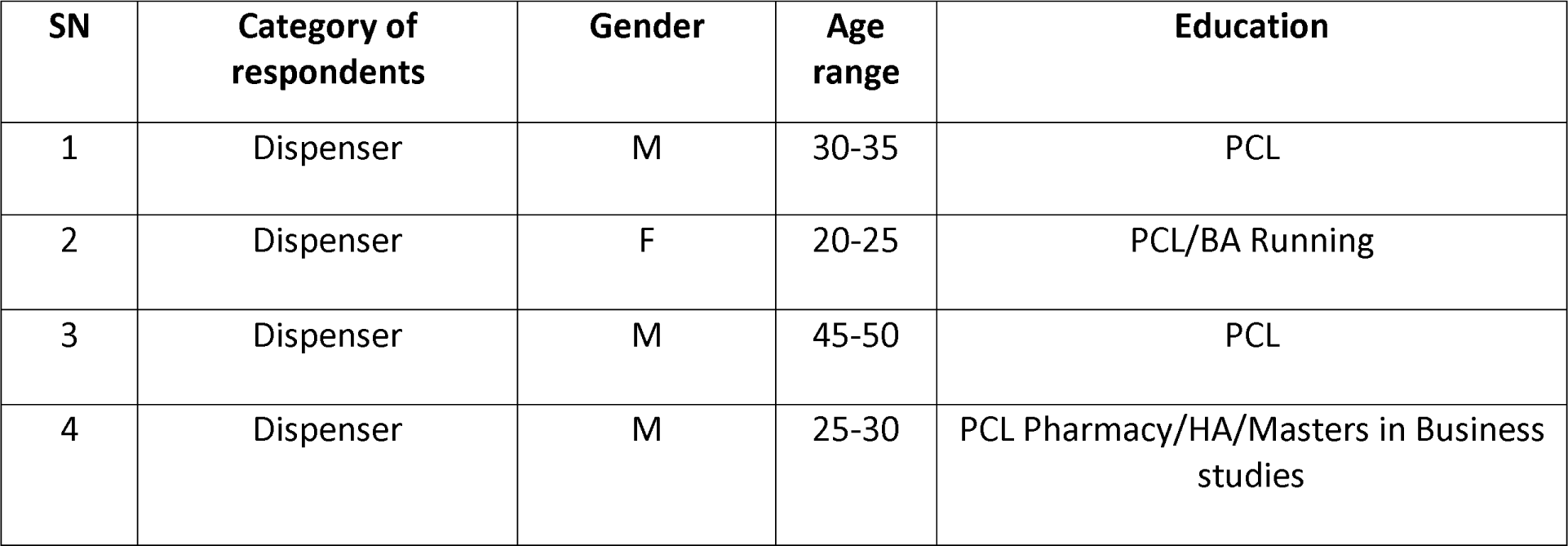
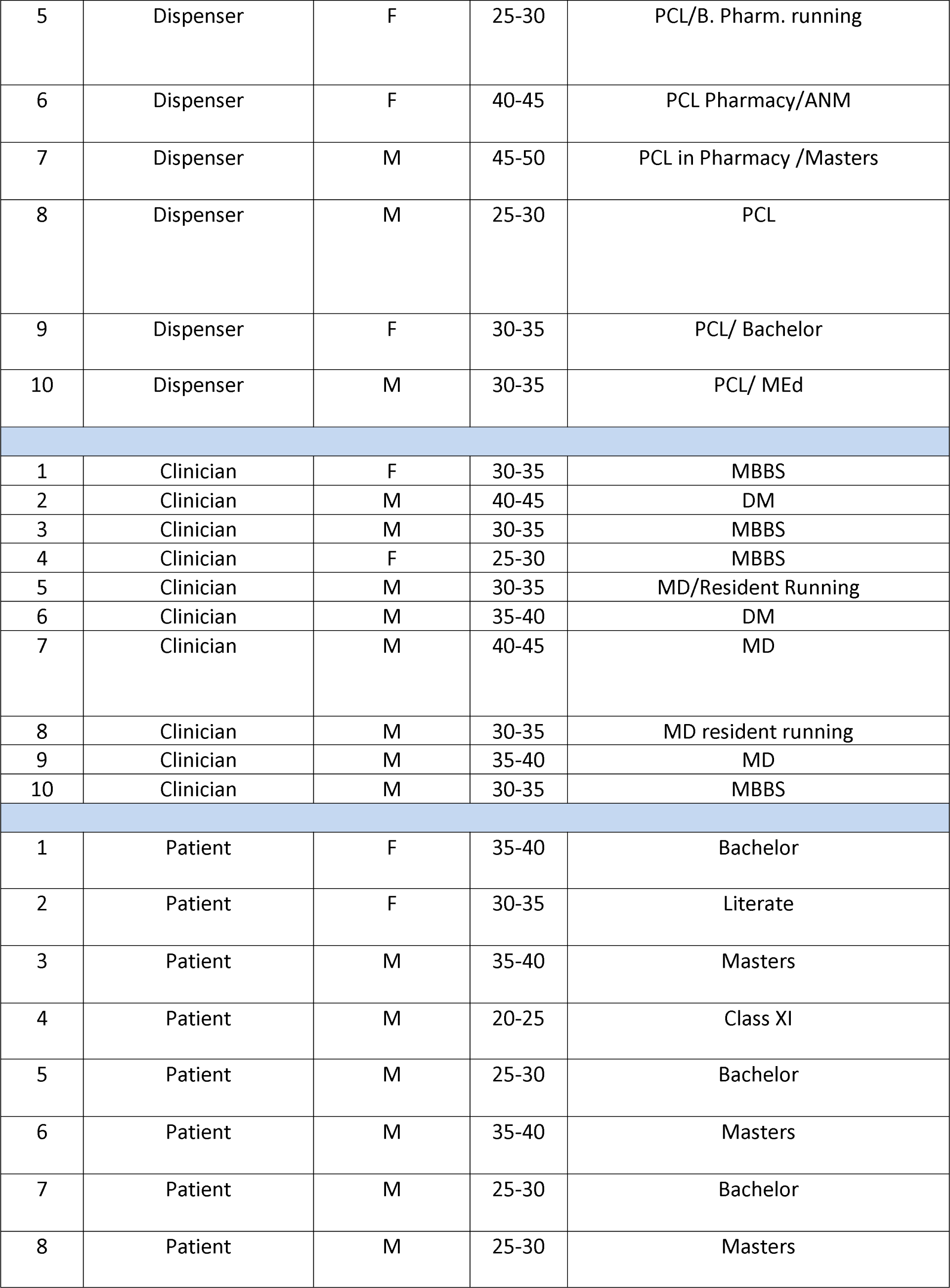
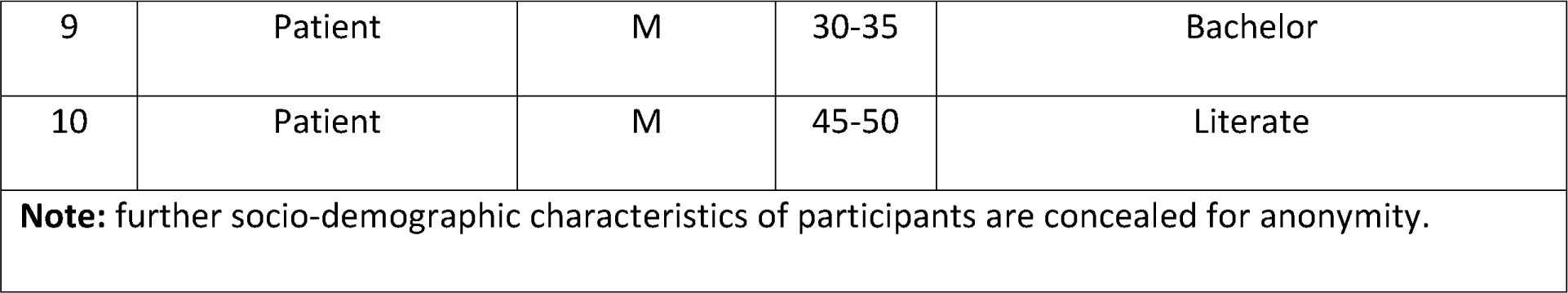
Socio-demographic characteristics of participants in the study

| SN | Category of respondents | Gender | Age range | Education |
| --- | --- | --- | --- | --- |
| 1 | Dispenser | M | 30-35 | PCL |
| 2 | Dispenser | F | 20-25 | PCL/BA Running |
| 3 | Dispenser | M | 45-50 | PCL |
| 4 | Dispenser | M | 25-30 | PCL Pharmacy/HA/Masters in Business studies |
| 5 | Dispenser | F | 25-30 | PCL/B. Pharm. running |
| 6 | Dispenser | F | 40-45 | PCL Pharmacy/ANM |
| 7 | Dispenser | M | 45-50 | PCL in Pharmacy /Masters |
| 8 | Dispenser | M | 25-30 | PCL |
| 9 | Dispenser | F | 30-35 | PCL/ Bachelor |
| 10 | Dispenser | M | 30-35 | PCL/ MEd |
| 1 | Clinician | F | 30-35 | MBBS |
| 2 | Clinician | M | 40-45 | DM |
| 3 | Clinician | M | 30-35 | MBBS |
| 4 | Clinician | F | 25-30 | MBBS |
| 5 | Clinician | M | 30-35 | MD/Resident Running |
| 6 | Clinician | M | 35-40 | DM |
| 7 | Clinician | M | 40-45 | MD |
| 8 | Clinician | M | 30-35 | MD resident running |
| 9 | Clinician | M | 35-40 | MD |
| 10 | Clinician | M | 30-35 | MBBS |
| 1 | Patient | F | 35-40 | Bachelor |
| 2 | Patient | F | 30-35 | Literate |
| 3 | Patient | M | 35-40 | Masters |
| 4 | Patient | M | 20-25 | Class XI |
| 5 | Patient | M | 25-30 | Bachelor |
| 6 | Patient | M | 35-40 | Masters |
| 7 | Patient | M | 25-30 | Bachelor |
| 8 | Patient | M | 25-30 | Masters |
| 9 | Patient | M | 30-35 | Bachelor |
| 10 | Patient | M | 45-50 | Literate |
| **Note:** further socio-demographic characteristics of participants are concealed for anonymity. |  |  |  |  |

### Data collection procedures

Two trained research assistants (BD and UTS; postgraduate researchers in Microbiology with skills in qualitative methods) conducted the SSIs in a quiet location chosen by respondents. SSIs were conducted based on the thematic guide designed by the study authors (**Supplementary File 2**). The investigators BA (MD, PhD) and KRR (MSc., PhD) trained the research assistants (BD and UTS) and initiated and supervised the procedures (such as obtaining of first few informed consents and execution of SSIs) on their physical presence. After an initial SSI of one person each from the desired stakeholder groups (clinicians, dispensers and patients), investigators (BA and KRR) held a reflection meeting to further instruct and train the assistants. The SSIs had the varying lengths of 13-42 minutes and were conducted in Nepali language.

The SSIs were conducted using a topic/subject guide tailored to each respondent group. The guides were developed by the research team and focused on: practices and frequency of prescription, sale and uses of antibiotics for COVID-19 patients, use of OTC antimicrobials and self-medication, basic awareness of AMR, and how the pandemic has impacted on the use of antibiotics and AMR. For interviews, all respondents were asked for a quiet location, without company. Respondents were interviewed at their workplaces with minimal distractions. For patients, interviews were conducted at the convenient locations recommended by them. All interviews were conducted face-to-face and were audio-recorded after obtaining the written informed consent forms. No interviews were repeated, and no participants were consulted for their transcripts.

### Data Analysis

Thematic analysis of the transcripts was conducted at various points of the study that entailed discussions of the findings and reflection among the interviewer (BD) and researchers (BA, KRR and SA). The transcripts and the findings were not shared among the respondents for their opinion, although some of the preliminary findings were re-discussed among the investigators and were prioritized in the subsequent interviews. Once the transcripts were ready, two researchers (BD and BA) had a line by line reading independently and were coded within the transcripts at first. The extensive report that included transcripts and interpretations were coded in QSR NVivo by BA and was cross-checked with the thematic synthesis by BD. Thematic analysis in this study fundamentally meant a use of deductive approach first to categorize the data based on the interview guides which were followed by addition of codes based on the line-by-line readings of the transcripts. All themes, both major and minor were discussed first among the data analysts (BA and BD) and then among the research team. Both major and minor themes identified from this study are the basis of the results in this study. Broadly, following previous qualitative work, data were triangulated from each of the respondents and are presented under broad themes. Excerpts were chosen based on their relevance to the themes in addition to the recurrences and uniqueness.

### Ethics approval

The study obtained ethical approval from Oxford Tropical Research Ethics Committee (OxTREC #502-22) and Nepal Health Research Council (Ref#156/2022).

## Results

### Overview of findings

Most of the respondents from all three groups in this study described increased use of OTC medications during the pandemic. A lot of the patients did not have knowledge about what constituted antibiotics and AMR. Except for a few dispensers, most of them did not understand the concept and mechanisms of AMR. Respondents perceived that antibiotics were able to reduce the morbidities and mortalities among COVID-19 patients. Patients perceived antibiotics as useful tools to tackle wide varieties of illnesses including the respiratory conditions that occurred during COVID-19. The fear and uncertainty attached to COVID-19 and the potential stigma if and when diagnosed made patients hesitant to visit formal health care services and instead resorted to informal health services. Hesitancy to visit the formal health care services also made patients to explore home remedies including wide spectrum of untested and unverified herbal products (**Figure 1**).

**Figure 1:**
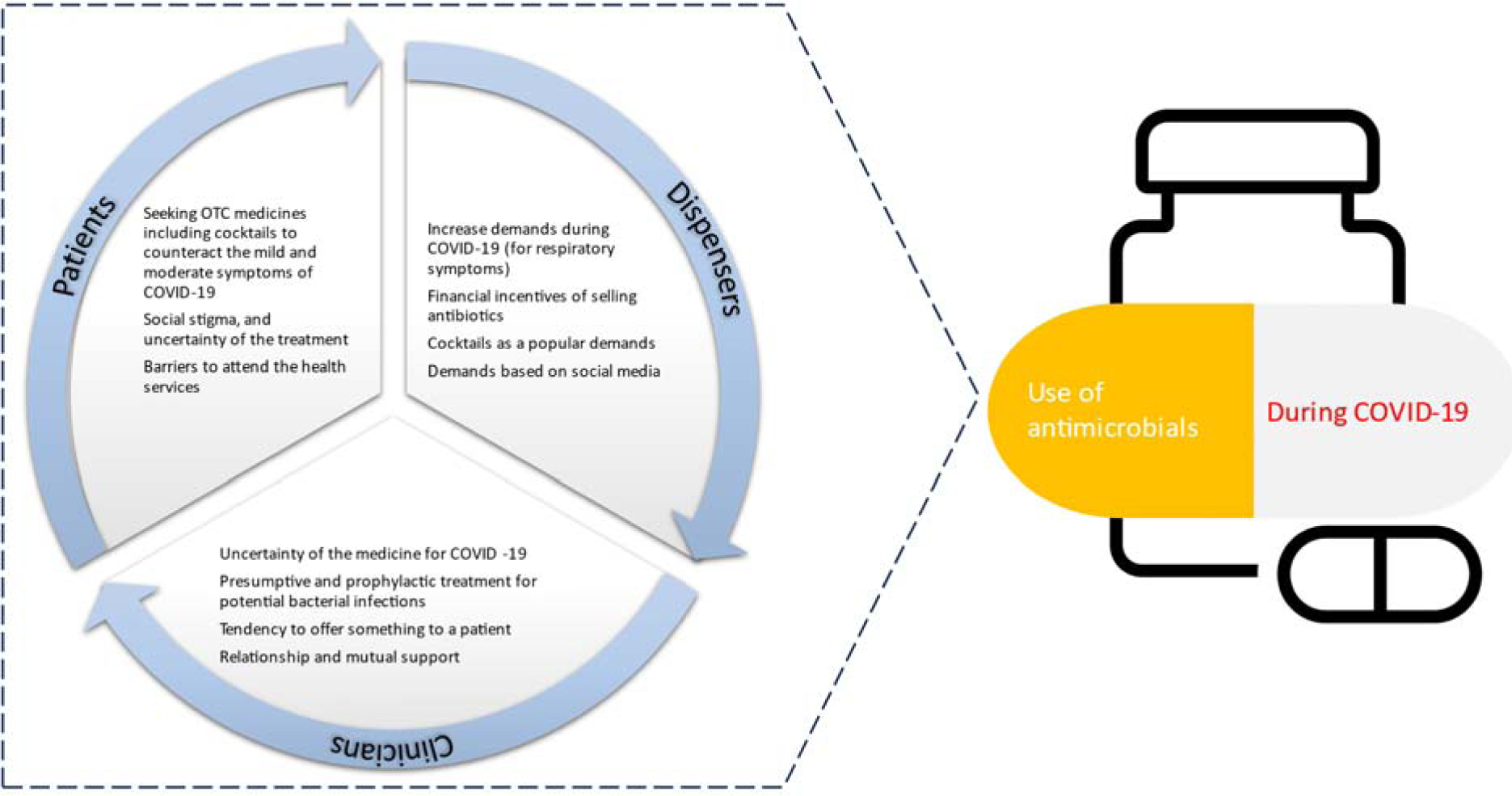
Thematic findings illustrating contributions to use of antimicrobials during COVID-19

### Treatment seeking and consumption of antibiotics during the pandemic

Patients shared their experience that they felt general weakness and discomforts even weeks or months after recovery from COVID-19. Respondents experienced severe symptoms while others had milder symptoms. Four patients in this study had to be admitted to high-dependency care units (HDUs) and intensive care units (ICU). Participants had suspected themselves of having been infected with COVID-19 infections even before they were confirmed by tests and sought all kinds of medical assistance.

Most of the patients followed home remedies with some traditional medicines (such as turmeric water, basil water, ginger water) for the first few days of symptoms onset. As the disease progressed, they sought assistance with OTC drugs such as paracetamol, multivitamins and antibiotics. Some patients were admitted to hospital just after three days of disease onset and had to be treated with oxygen support. Respondents visited labs mostly when they suspected that the illness could be COVID-19 and could be transmitted to their family members. Patients also visited labs when they failed to resolve their conditions after self-medication including trying wide range of home remedies. Patients shared that most of their suspected close-contacts and acquaintances wanted to evade the laboratory diagnosis of the COVID-19 because of fear of mandatory isolation, social stigma (especially during the first wave in which people used to think of the novel virus as a mysterious agent of the disease), and lack of definitive treatment. Patients explained that even when they were diagnosed as COVID-19, they would not have any treatment available which discouraged them to diagnose their illness. The lack of definitive treatment (with or without diagnosis) also led patients to follow some random combinations of home remedies and OTC medicine including antibiotics.

*Most of the people wanted to avoid testing because they were in constant fear that if somebody knows their infected status, they could be given a bad name by the society, Koroné here as an instance as it used to be with TB and leprosy patients when I was a child. Not only the patients, but also their entire family members used to be boycotted and cursed by society during that period. Even the succeeding generations had to endure a bad name [koriko santaan here as an instance; nodular lesions of leprosy was called as Kor in vernacular language and koriko santaan was name given to the blood/offspring of a leprosy patient]. The generation which had witnessed and endured such discrimination and stigma attached with the disease, avoided themselves to test or confess their illnesses*.

**45-50 years old, male, patient**

Most patients did not know what antibiotics really were, although they did hear about antibiotics and some perceived that antibiotics were one of the strong medicines. Many patients recalled the drug Azithromycin which they had used as OTC medicine during the pandemic, although most did not know if this was an antibiotic. Patients were unable to recall the names of the antibiotics because they were unaware of the other names and many confirmed that Azithromycin was one of the most bought medicines during the pandemic. Other antibiotics were also used (prescribed by dispensers and doctors) but patients could not recall their name.

The patients had a habit of buying antibiotics OTC for themselves and family members. Most of them had habits of purchasing it 2-3 times a year. Most of the patients thought that antibiotics were useful, often lifesaving based on their understanding of the term ‘antibiotics’. However, some of the respondents said that they would never buy OTC drugs. This awareness stemmed from the unfavorable experiences they have had during the course of COVID infection. The utility of antibiotics was thought to be high for the patients with severe symptoms while there was little or no use for those with milder symptoms. There was a mixed reaction on the use of antibiotics before and after the pandemic while most of them agreed that there was increased use of antibiotics during the peaks of pandemic. Most of them thought that AMR might have increased due to pandemic.

Participants also linked using OTC drugs to their hesitancy to visit large private health care institutions as these institutions were deemed to be commercially motivated which essentially meant that participants were prescribed various diagnostics and medications, sometimes unnecessarily. The diagnostics and number of medications were perceived to be a burden to patients, and thus resorted to evading formal health services by visiting the drug shops. Nonetheless, buying OTC medication was realized to be sub-optimal for their care.

*There could be both benefits and harms. Benefit from the perspective of short-term relief but in the long-term the OTC medicines may bring out several health hazards. Nowadays doctors are waiting patients so as to prescribe and sell more and more drugs to increase their business volumes. This sector has been commercialized by not only pharmacies but also by doctors themselves. Even if a patient does not need antibiotics, they want to prescribe and sell their desired products. Doctors and healthcare facilities today are very far from those who do not have a good income. I could not find any hospital for admission during the pandemic time although I made phone calls to several hospitals in the valley*.

**35-40 years old, male, patient**

Patients also shared their pre-existing habits of seeking OTC medications for the common symptoms such as fever, cold, tonsilitis, sore throats and wounds. Patients also shared that they used to buy an entire dose of antibiotics at once and rarely completed the dose of purchased antibiotics; they stopped taking it once they began to feel comfortable (subsiding of the symptoms); and rarely visited pharmacies or hospitals for follow up. Most of the patients revealed that they abandoned the dose after the first three days of taking the drug.

*I buy the entire dosage at once. I rarely go for follow-up; if I am feeling better, why should I?*

**25-30 years old, female, patient**

### Antibiotics selling behavior

When asked about the notable changes dispensers had observed during their services, most of them appreciated the advancement in healthcare facilities, improvement in accessibility and raised awareness towards individual and family health. However, dispensers at the same time were worried about the progressive ineffectiveness of the antibiotic (and other) drugs. Such progressive ineffectiveness of the antibiotics was attributed to self-medication and demands with specific medications from patients.

*There is a drastic change. During the initial days of my service, a low-dose simple combination used to be sufficient for most of the complaints, which is of no use these days. Patients used to follow our guidelines and suggestions. Nowadays, people [either] are well-educated or pretend to be learned. I do not know, but it is difficult to convince them [these days]. A sense of distrust towards healthcare service providers is well observed in present days*.

**20-25 years old, female, dispenser**

When discussed about the drivers of OTC medication, the major source of information were various websites surfed through Google and social media.

*They [customers/visitors/patients] are following close-contacts, social media, YouTube and Google search medium rather than specialists. They have more belief in those media than us. I do not say these media are not good, but how would they identify the true information when everything is flooded there? Referring to the examples of someone who has recovered from any specific symptom with a specific drug and exerting pressure for the same drug when similar symptoms are developed has become a new normal these days*.

**40-45 years old, male, dispenser**

Dispensers were also concerned about the growing distrust of the people towards healthcare workers and the government. Distrust towards the dispensers were attributed to perception that the healthcare workers were mostly motivated by monetary incentives than the wellbeing of the patients. Such distrust was thought to be the main drivers towards OTC medications which apparently helped patients to bypass the formal health care workers and the service institutions.

Dispensers agreed that they had prescribed antibiotics mostly based on their presumptive diagnosis rather than the diagnosis based on the lab confirmation. During the pandemic, dispensers agreed that there was an increase in the sales of antipyretics, analgesics, multivitamins, and antibiotics as major drugs to subside the symptoms of COVID. However, there were mixed reactions on the overall sales of antibiotics during the pandemic. Dispensers also reported that there were no significant increase in number of visitors during the first wave of the pandemic but there was a substantial increase during second and third waves.

Nonetheless, dispensers agreed that they had received increased drug-pressures from the patients during COVID pandemic compared to pre-COVID times. Related to the type of patients in terms of demanding the antibiotics, dispensers shared that the seemingly educated ones generally dictated the demands while illiterate patients generally came with previous drug leaflets and empty blister package to demand for exact same medications, most of which were antibiotics. On some occasions, patients demanded the antibiotics referring to the need for their family, friends and relatives to exert indirect selling pressure. There was an increasing trend of OTC drugs and selling pressures on these dispensers.

Based on the dispensers’ opinion, respiratory and diarrheal illnesses were the major reasons for sales of antibiotics. Driven by excessive drug-pressure and self-prescription (medication), dispensers did not reject selling those drugs sought by patients although they they tried convincing patients not to opt for antibiotics at the first step.

*We would try to figure out the person psychologically. If he seems to be convincing, of course we would. But if the person seems to be stubborn and even on the verge of switching my pharmacy [in case he does not get his desired drugs], we simply deliver our intended message in not so stressing manner. After all, it is our business too. Who would like to lose a valued customer?*

**40-45 years old, female, dispenser**

The respondent further added: *People are mostly preoccupied. Even if the handwriting of the doctor is not well understood and even if there were availability of quality drugs from another company, people would not like to listen. They hear only what they want to hear. They [would] rather visit a dozen pharmacies for their drug of choice but would hardly get convinced*.

Apart from inappropriate drug pressures, irrational doses were of another concern. Some patients used to ask for just a couple of tablets of antibiotics, and denying such requests were out of scope for dispensers as patients would switch to generally an adjacent drug store.

Dispensers from community pharmacies experienced the OTC sales of antibiotics constituting more than two-third of the total sales of antibiotics volume. In general dispensers did not have their lab for confirmatory diagnosis, and thus had to resort to empirical dispensing of the antibiotics. The antibiotics most often dispensed included azithromycin, amoxicillin, cefixime, ampicillin, levofloxacin and clavulanic acid during and after the pandemic. Dispensers believed that these antibiotics were the most extensively used drugs in terms of covering most of the infectious diseases that they have encountered so far. Dispensers did not reject selling OTC antibiotics although some dispensers of hospital pharmacies revealed that they recently began rejecting it because of the fear from regulatory bodies.

*We have recently begun to reject some of the requests. It’s mainly because of fear from authorities as there are stringent regulations being implemented these days*.

**25-30 years old, female, dispenser**

Most of the drug dispensers had no specific knowledge regarding AMR and the public health crisis due to its emergence. When asked about the knowledge and cautionary measures required to counter the COVID-19, respondents were aware of the symptoms, disease manifestations and precautions to be taken while dealing with the infected individuals. OTC drugs were offered to patients based on the symptoms that resembled COVID-19. Regarding the need and effectiveness of antibiotics in the management of COVID-19, the respondents believed that antibiotics had impact on COVID-19 but did not know the specific mechanisms, how it cured the infection. In terms of timing, dispensers believed that patients attended their drug shops once their home remedies failed to cure.

Dispensers also believed that antibiotics were expensive but despite so, demands and particularly the demands for incomplete dosage were problematic, which was not just motivated by the potential revenues by selling the full course, but they saw the adversities of underdosing. The excessive use of antibiotics, and the incomplete dosing could contribute to the existing problem of AMR. Irrespective of their own shortcomings, dispensers knew and expressed that OTC use of antibiotics needed scrutinization, and wanted to convey message to patients.

*I request you all the patients to avoid quackery (pretension of well understanding of antibiotics among educated and mass people during these days) and listen to the trained healthcare professional. Further I stress on the need for a completion of a full course of treatment among the patients undergoing antibiotic therapy*.

**45-50 years old, male, dispenser**

### Practices regarding antibiotics prescription

When asked about the specific changes before and after the pandemic, most of the clinicians witnessed increased respiratory complaints and other post-COVID illnesses including psychological problems (anxiety, increased heartbeat, and panic attack) and heart diseases. Clinicians were aware of OTC use of antibiotics, AMR and their potential consequences. A significant majority of the clinicians had experienced drug pressure from patients. Even an intensivist shared his experience as

*Majority of the patients ask drugs [basically painkillers and some antibiotics] for themselves complaining of the pains, discomforts or other illnesses. However, we never prescribe unnecessary drugs. We tell the patients that the drugs they intend to consume may not be necessary and useful rather may be harmful. We do not get them convinced*.

**40-45 years old, male, clinician**

Clinicians listed the following symptoms as the major drivers behind OTC sales of antibiotics: respiratory tract infections (sore throat, common cold cough, and tonsillitis), fever and urinary tract infections. These symptoms were similar to that of COVID-19 which might have led to increased use of OTC drugs during the pandemic. Resemblance of the COVID-19 symptoms to the known conditions such as respiratory infections also in fact promoted OTC use of antibiotics because of the seemingly familiar symptoms which were generally cured by the OTC drugs in the past. Second and third waves were deemed to mount pressure among clinicians as well to prescribe antibiotics. The scenario was intense in rural regions where clinicians were under pressure to offer antibiotics because the patients travelled from far to attend the health facilities. These patients were unlikely to follow-up for monitoring of symptoms, and took complacence through the antibiotics irrespective of their need and impact on their illnesses. Sometimes clinicians had to prescribe antibiotics to avoid the potential hostility with patients resulting from sending patients empty handed.

*I think it [handling pressure from patients] mainly depends on the physician’s side. If one can convince the patients, probably they would likely understand. Some proportion (around 10%) would still be not convinced. If the physician pays attention and spares ample time, the patient would likely be convinced but the problem with us is that we cannot give enough time and counseling [due to the high number of visitors per physician]. And some physicians are taking this reason [default condition] as a means of excuse even when they have enough time. In rural settings, sometimes physicians in far off districts do not want to displease the locals, as physicians have to get familiar with them so that they can continue their job smoothly. Who wants to confront the locals even if they are constantly pressurizing for a specific drug?*

**30-35 years old, male, clinician**

Paracetamol and pain killers, anti-diarrheal, antibiotics, salbutamol for breathlessness, and other topical antibiotics for wounds and other skin infections were recalled as the major drugs being asked by the patients. Unsurprisingly, most of these drugs were common OTC medications all over the country.

*I can tell you with certainty that asking for antibiotics and OTC use of antibiotics is rampant all over the country. Amoxicillin, ciprofloxacin are the chief used antibiotics in rural settings as it is provided freely by the government and are common medicines for villagers*.

**40-45 years old, male, clinician**

Some physicians expressed worries regarding the drug pressure they used to receive from well-educated patients. They used to exert pressure referring to the information they accessed over the non-specific web search engines (such as Google) of the internet. Some experienced an increased pressure of drugs in post-COVID time referring to the patients’ altered behavior in seeking medical care.

*It has increased in post-COVID time. It may be due to the fact that before pandemic patients could access hospital facilities easily. However, during the pandemic, this easy access was greatly reduced due to lockdown and other travel restrictions which compelled them to rely on pharmacies available nearby. In this way, pharmacies may have prescribed and/or sold unnecessary drugs as a lucrative means which may have altered the patients’ behavior in seeking medical care and purchasing drugs*.

**35-40 years old, male, clinician**

Physicians were prescribing antibiotics daily as an essential aspect of their treatment therapy. Azithromycin, amoxycillin, cefixime, ampicillin, levofloxacin, amoxiclav, cephalosporin, and ofloxacin/ciprofloxacin were frequently prescribed antibiotics by physicians. These drugs were the most used OTC antibiotics in the country. Participants recognized azithromycin as the most sold antibiotic in the country while some of them listed azithromycin and amoxicillin as comparable drugs in view of business volume.

When asked about the antibiotic prescriptions, clinicians resorted largely to empirical practices referring to the reasons that most of the patients were already having antibiotics from OTC and when symptoms did not subside visited their clinic. This particular patients’ behavior interfered the diagnosis, as culture and drug susceptibility tests were bound to be altered by the OTC use of antibiotics. Thus, clinicians prescribed higher generation antibiotics to counteract the unresolved infections. Clinicians also shared that most patients who visited the clinic came from remote and rural regions and did not have time, money and patience to wait for the culture results which also obliged clinicians to be context friendly by offering empirical antibiotics at an OPD basis. Clinicians also used antibiotics based on the length and severity of the infection.

*Duration and severity of the onset of the symptoms play a crucial role in our response. If the duration is shorter, we don’t go directly with antibiotic treatment rather we aim for symptomatic management. However, if there is [an] increase in duration and severity, we have to go for antibiotic therapy. It’s again empirical than following any specific protocol*.

**30-35 years old, female,**

Clinicians had experienced the poor follow-up among patients who were prescribed antibiotics. Most patients in their experience did not complete the full course of treatment. Patients also brought the old prescriptions, empty blister packs and made request for the prescription. As most of the clinicians had past experiences of service in the peripheral/rural part of the country, there was a unanimous view that OTC drugs (including antibiotics) were prevalent in any corner of the country. They expressed that ‘*empirical prescription will not be replaced anytime soon, given the lack of diagnostic facilities in the majority of the healthcare centers that covers about two-third of the total population.’*

Although the doctors believed that prescribing antibiotics were one of the counter strategy to tackle the disease progression (e.g. secondary bacterial infections), they did not consider the therapy at first. Clinicians were well aware that antibiotics had nothing to do with the viral infections including COVID-19. But as the disease progressed, secondary bacterial infections were prevalent among severely ill patients. Antibiotics were therefore inevitably prescribed to prevent secondary bacterial infections including when they had definitive diagnosis of bacterial infections. Antibiotics were considered a lifesaving option. Nonetheless, clinicians echoed with both patients and dispensers that self-medication and subsequent drug pressure for OTC was more prevalent among patients with milder symptoms than the ones who were admitted. Clinicians shared some of the most prescribed antibiotics during the pandemic included azithromycin, piperacillin/tazobactam and cephalosporins among the patients who were admitted at the ICU.

When doctors were asked about the future applicability of antibiotics in COVID-19, they could not rule out the drug’s necessity among COVID patients with co-morbidities and other potential bacterial (secondary and nosocomial) infections during the course of infection. Physicians were aware about the drug resistance, and believed that AMR may have been flared up due to excessive and inappropriate use of antibiotics during the pandemic. Clinicians shared serious concerns about the potential consequences that can result from AMR and reflected on their own roles and obligation to combat burgeoning surge of AMR.

*Nowadays, we admit patient in ICUs and request for the antibiotic sensitivity test where we encounter the resistance to almost all of the available drugs. Who is responsible for this? Definitely we are. We physicians are mainly responsible for this situation. We are taught to escalate rather than to deescalate the situation [sarcasm]. We are much poorer when it comes to antibiotic stewardship. We merely follow our seniors rather than questioning the long-held practices. There is over consciousness, lack of knowledge and limitation in our diagnostic capacities [which is] a major problem. But we should take the responsibility from our side rather than blame. And from the policy level side, it is seriously affected by various problems. These are the situations that have prevailed among us*.

**30-35 years old, male, clinician**

## Discussion

This is the first study in Nepal which attempted to document the use of antibiotics during the pandemic from the experiences of COVID patients, dispensers and clinicians. The use of OTC antibiotics was accentuated during the COVID-19 pandemic and several reasons for such an increased use of antibiotics were found. Both empirical treatment through prescription from clinicians and OTC based use of antibiotics remained high during the COVID-19 pandemic which echoes with the previous studies from Scotland [24], Saudi Arabia [25], Peru [26], and Bangladesh [27]. The overuse of antibiotics during the pandemic has been echoed globally [11]. COVID-19 was deemed to be a special circumstance that added extraordinary pressure on both physicians and drug dispensers [24]. Although clinicians perceived the need to halt the severe infections and prevent the potential secondary infections following COVID-19, dispensers’ incentives for OTC sales of antibiotics were primarily patients’ demands. This finding is analogous to reports from Eastern Mediterranean regions [28] and previous reports from Nepal [29-31]. Although patients did agree that they may have visited dispensaries more frequently, a lot of these patients were also unaware of what constituted antibiotics and the meaning and consequences of AMR. COVID-19 created a facilitative background for all stakeholders to overuse antibiotics. Nonetheless, the use of antibiotics through the dispensary outlets remains to be sub-optimal and detrimental because they were also underdosed and were based on the non-expert (patients’) demands, and dispensers’ incentives.

Aligning with the previous studies [30, 31], respondents’ knowledge on antibiotics were poor and thought these were a powerful group of drugs and referred to as a ‘big boss’, a ‘final solution’ or a ‘different class of high dose Superdrug’, and also echoes with the study from Bangladesh [32]. Although dispensers did have knowledge about antibiotics, they did not understand the concept and mechanisms of AMR. Inevitably, very few patients knew what antibiotics were and AMR was in general a complex concept.

## OTC use of antibiotics during COVID-19

Most of the patients wanted to avoid the confirmatory diagnosis because of the social stigma attached to COVID-19, specifically, fear of social isolation, mandatory quarantine, and also the lack of definitive treatment for it. Some of these drivers were echoed by previous study from Nepal [33] while a study from 28 countries documented social stigma as the most important factor in contributing the perceived risk and fears as the root cause of averting health-seeking behaviors and practices at formal health services [34-38]. The other reasons to avoid seeking definitive diagnosis and treatment were because patients considered COVID symptoms as simple case of ‘common cold’ because of the resemblance and resorted to OTC antibiotics.

Patients in this study followed home remedies and OTC medicines before they sought medical attention in hospitals. Our finding has been corroborated by several other studies from Asian and African countries [39-41]. Private pharmacies and dispensers comprised the larger chunk of informal prescription and sales of OTC medicines to these patients; this finding echoes with several previous reports [42-44]. Other reasons behind this informality were patients’ self-medications and induced pressure on dispensers, often guided by their close-contact and social media. Similar compelling factors were also outlined in the previous studies [15, 45]. Home remedies comprised mainly water boiled with basil leaves, ginger and turmeric together or separately as per their personal preferences. This combination has been a long-held tradition in Nepal where Ayurveda and homeopathy has been a source of treatment for febrile illnesses including flu-like, and common colds—symptomatically similar to COVID-19. Such traditions were prevalent in various settings across the globe [39-41, 46]. Most of the patients shared a feeling of lowered self-esteem and psychological crises in the course of infection as seen in other studies [35]. Apart from the home remedies, building self-confidence and thus immunity was also promoted among peers and in social media platforms, nonetheless the promotions were also unevidenced and tended to recommend wide variety of medications including herbs and unlicensed products [47].

Dispensers also echoed on heightened use of antibiotics during the pandemic. Although the major reasons for high use of antibiotics were attributed to patients’ demands during non-COVID times, their default position on selling antibiotics were inundated by the claims and demands they believed were mostly driven by social media and what they heard from their relatives, neighbors and friends [15, 45, 48]. The uncertainty of the pandemic in terms of what COVID is, how it is caused, and the potential causative agents are known contributors to the OTC havoc and follows a global trend [27]. OTC demands for antibiotics were a pre-existing trend and behavior in Nepal [20, 30, 31] and were accentuated by the added complexity of COVID-19. Worryingly, these demands had several flaws in itself that for example included buying of incomplete doses and poor adherence to the antibiotics when they needed it [49, 50]. Although dispensers’ acceptance of increased OTC sales of antibiotics is clear indication of how pandemic accentuated the inappropriate use of antibiotics, dispensers were restrained by their commercial interests in not losing the customers by complying with the demands [49, 51]. Knowledge that sales of antibiotics without prescriptions and potential adversities was understood to some extent by the dispensers, but the policy restrictions and ethical good practice were beyond their scope when it came to daily practice. Nonetheless, during interactions, dispensers also attempted to display their knowledge about the adversities of dispensing antibiotics even if the practice did not approve their opinion.

Clinicians echoed with both patients and dispensers that there was an accentuated use of antibiotics during the pandemic and the pressure on prescribing antibiotics were felt by the clinicians. Two kinds of pressures were explained by the clinicians, one that was intrinsic inherent in their own practice and another one was extrinsic that emerged from patients and their relatives. These findings were also supported by some previous studies [24, 52-55]. COVID-19 manifested as respiratory illnesses and there was an increase influx of patients who either already had OTC antibiotics or were brought severely ill at the tertiary care centers. The obvious delays in the diagnosis of the causative agents (such as bacterial infections) meant that clinicians had to resort to empirical treatment to prevent the secondary infections [6, 56]. Also, the increase in respiratory symptoms compelled clinicians to offer antibiotics to prevent its further progression. In addition, clinician’s practice to prescribe or offer something to a patient presenting with respiratory illness is also socially woven to be seen as kind and ‘giving’ rather than being strict on prescribing and adding disappointments to patients [24]. Antibiotics prescription for patients were also a symbolic gesture of social niceties, relationship and trust. Especially, during the pandemic where trust towards health care workers was endangered due to uncertainty, antibiotics may have had an impact as a social binder, a commonest denominator contributing to the acceptance of health interventions [47, 57].

## Strengths and limitations

Respondents (mostly dispensers and clinicians) in this study may have been affected by the social desirability bias. Thus, their views and perspectives might be different from their practice, nonetheless, follow-up and indirect questions were asked to report the overall behavior of stakeholders that may have offered space to discuss the topics more openly without self-attributing. Some of the respondents, especially dispensers, tried to avoid the interviewer because of the qualms and suspicion that they may have been monitored or inspected by authorities or journalists. Although interviewer attempted to balance the gender among respondents, higher proportion of clinicians were male reflecting the sex ratio among the doctors’ population in Nepal.

## Conclusions

COVID-19 exerted an extraordinary pressure on the use of antibiotics in Nepal. Both prescription and non-prescription (OTC) use of antimicrobials were found high during the pandemic. COVID-19 was deemed to be a special circumstance that added extraordinary pressure and demands on both physicians and drug dispensers. While clinicians prescribed antibiotics to arrest the severity of infection, prevent secondary infection and to maintain the patient’s demands, dispensers’ selling of antibiotics was mostly motivated by their own commercial interests, and patients’ demands. The increase in OTC sales of antibiotics was also inappropriate because of the sale of incomplete dosage/duration. Regulatory policies on antibiotics need a revised contingency plan specifically to tailor for special circumstances such as COVID-19.

## Supporting information

## Data Availability

The data is available upon request to the Mahidol Oxford Tropical Medicine Research Unit Data Access Committee (http://www.tropmedres.ac/data-sharing) complying with the data access policy (http://www.tropmedres.ac/_asset/file/data-sharing-policy-v1-0.pdf).

## Acknowledgements

We extend our thoughtful gratitude to all the participants who were involved in this study.

## Author Contributions

Conceptualization: BA, PYC, CP, DL, AK, SP; Investigation: BD; Data collection, and transcription: BD, AB, UTS; Data curation: BD, BA; Formal analysis: BA, BD; Methodology: BA, BD; Project administration: SA, KRR, NA; Supervision: BA, PYC, CP, KRR, PG; Visualization: BA; Writing-original draft: BA, BD; Writing-review and editing: all authors

## Funding

The study was funded by Wellcome Trust, UK. For the purpose of Open Access, the author has applied for a CC BY public copyright license to any Author Accepted Manuscript version arising from this submission. The funders had no role in study design, data collection and analysis, decision to publish, or preparation of the manuscript.

Supplementary File 1: COREQ (Consolidated criteria for Reporting Qualitative research) guideline.

Supplementary File 2: SSI thematic guide for dispensers, patients and clinicians

## References

1. WHO. WHO Coronavirus (COVID-19) Dashboard [May 13, 2023]. Available from: https://covid19.who.int/

2. Krammer F. SARS-CoV-2 vaccines in development. Nature. 2020;586(7830):516-27.

3. Winch GM, Cao D, Maytorena-Sanchez E, Pinto J, Sergeeva N, Zhang S. Operation Warp Speed: Projects responding to the COVID-19 pandemic. Project Leadership and Society. 2021;2:100019.

4. Subramanya SH, Czyż DM, Acharya KP, Humphreys H. The potential impact of the COVID-19 pandemic on antimicrobial resistance and antibiotic stewardship. Virusdisease. 2021;32(2):330-7. Epub 20210525. doi: 10.1007/s13337-021-00695-2. PubMed PMID: 34056051; PubMed Central PMCID: PMCPMC8145182.

5. O’neill J. Antimicrobial resistance: tackling a crisis for the health and wealth of nations. Rev Antimicrob Resist. 2014.

6. Seneghini M, Rüfenacht S, Babouee-Flury B, Flury D, Schlegel M, Kuster SP, et al. It is complicated: Potential short- and long-term impact of coronavirus disease 2019 (COVID-19) on antimicrobial resistance-An expert review. Antimicrob Steward Healthc Epidemiol. 2022;2(1):e27. Epub 20220218. doi: 10.1017/ash.2022.10. PubMed PMID: 36310817; PubMed Central PMCID: PMCPMC9614949.

7. Aslam A, Gajdács M, Zin CS, Ab Rahman NS, Ahmed SI, Zafar MZ, et al. Evidence of the practice of self-medication with antibiotics among the lay public in low-and middle-income countries: a scoping review. Antibiotics. 2020;9(9):597.

8. Nepal G, Bhatta S. Self-medication with antibiotics in WHO Southeast Asian Region: a systematic review. Cureus. 2018;10(4).

9. Pokharel S, Adhikari B. Antimicrobial resistance and over the counter use of drugs in Nepal. J Glob Health. 2020;10(1):010360. Epub 2020/06/23. doi: 10.7189/jogh.10.010360. PubMed PMID: 32566152; PubMed Central PMCID: PMCPMC7296207 Interest form (available on request from the corresponding author) and declare no competing interests.

10. Alshaikh FS, Godman B, Sindi ON, Seaton RA, Kurdi A. Prevalence of bacterial coinfection and patterns of antibiotics prescribing in patients with COVID-19: A systematic review and meta-analysis. PLoS One. 2022;17(8):e0272375.

11. Nandi A, Pecetta S, Bloom DE. Global antibiotic use during the COVID-19 pandemic: Analysis of pharmaceutical sales data from 71 countries, 2020–2022. Eclinicalmedicine. 2023;57.

12. Haldane V, De Foo C, Abdalla SM, Jung AS, Tan M, Wu S, et al. Health systems resilience in managing the COVID-19 pandemic: lessons from 28 countries. Nat Med. 2021;27(6):964–80. Epub 20210517. doi: 10.1038/s41591-021-01381-y. PubMed PMID: 34002090.

13. Shrestha N, Mishra SR, Ghimire S, Gyawali B, Marahatta SB, Maskey S, et al. Health system preparedness for COVID-19 and its impacts on frontline health care workers in Nepal: a qualitative study among frontline healthcare workers and policymakers. Disaster Med Public Health Prep. 2021:1–29. Epub 2021/06/19. doi: 10.1017/dmp.2021.204. PubMed PMID: 34140051.

14. Mazzon D. [Ethical use of antibiotics in the era of multiresistance: a common good for the individual or the society?]. Recenti Prog Med. 2016;107(2):71–4. doi: 10.1701/2152.23268. PubMed PMID: 26901582.

15. Wegbom AI, Edet CK, Raimi O, Fagbamigbe AF, Kiri VA. Self-Medication Practices and Associated Factors in the Prevention and/or Treatment of COVID-19 Virus: A Population-Based Survey in Nigeria. Frontiers in public health. 2021;9:606801. Epub 20210604. doi: 10.3389/fpubh.2021.606801. PubMed PMID: 34150693; PubMed Central PMCID: PMCPMC8213209.

16. Langford BJ, So M, Raybardhan S, Leung V, Soucy JR, Westwood D, et al. Antibiotic prescribing in patients with COVID-19: rapid review and meta-analysis. Clin Microbiol Infect. 2021;27(4):520–31. Epub 20210105. doi: 10.1016/j.cmi.2020.12.018. PubMed PMID: 33418017; PubMed Central PMCID: PMCPMC7785281.

17. Torres NF, Chibi B, Middleton LE, Solomon VP, Mashamba-Thompson TP. Evidence of factors influencing self-medication with antibiotics in low and middle-income countries: a systematic scoping review. Public Health. 2019;168:92–101. Epub 20190201. doi: 10.1016/j.puhe.2018.11.018. PubMed PMID: 30716570.

18. Do NTT, Vu HTL, Nguyen CTK, Punpuing S, Khan WA, Gyapong M, et al. Community-based antibiotic access and use in six low-income and middle-income countries: a mixed-method approach. The Lancet Global health. 2021;9(5):e610–e9. Epub 20210310. doi: 10.1016/s2214-109x(21)00024-3. PubMed PMID: 33713630; PubMed Central PMCID: PMCPMC8050200.

19. Molento MB. COVID-19 and the rush for self-medication and self-dosing with ivermectin: A word of caution. One Health. 2020;10:100148. Epub 20200624. doi: 10.1016/j.onehlt.2020.100148. PubMed PMID: 32632377; PubMed Central PMCID: PMCPMC7313521.

20. Pokharel S, Raut S, Adhikari B. Tackling antimicrobial resistance in low-income and middle-income countries. BMJ Glob Health. 2019;4(6):e002104. Epub 2019/12/05. doi: 10.1136/bmjgh-2019-002104. PubMed PMID: 31799007; PubMed Central PMCID: PMCPMC6861125.

21. Rizvi SG, Ahammad SZ. COVID-19 and antimicrobial resistance: A cross-study. Sci Total Environ. 2022;807(Pt 2):150873. Epub 20211008. doi: 10.1016/j.scitotenv.2021.150873. PubMed PMID: 34634340; PubMed Central PMCID: PMCPMC8500695.

22. Rayamajhee B, Pokhrel A, Syangtan G, Khadka S, Lama B, Rawal LB, et al. How well the government of Nepal is responding to COVID-19? An experience from a resource-limited country to confront unprecedented pandemic. Frontiers in public health. 2021;9:597808.

23. Saunders B, Sim J, Kingstone T, Baker S, Waterfield J, Bartlam B, et al. Saturation in qualitative research: exploring its conceptualization and operationalization. Qual Quant. 2018;52(4):1893–907. Epub 2018/06/26. doi: 10.1007/s11135-017-0574-8. PubMed PMID: 29937585; PubMed Central PMCID: PMCPMC5993836.

24. Duncan EM, Goulao B, Clarkson J, Young L, Ramsay CR. ’You had to do something’: prescribing antibiotics in Scotland during the COVID-19 pandemic restrictions and remobilisation. British dental journal. 2021:1–6. Epub 20211123. doi: 10.1038/s41415-021-3621-8. PubMed PMID: 34815483; PubMed Central PMCID: PMCPMC8609985.

25. Khojah HMJ. Over-the-counter sale of antibiotics during COVID-19 outbreak by community pharmacies in Saudi Arabia: a simulated client study. BMC Health Serv Res. 2022;22(1):123. Epub 20220129. doi: 10.1186/s12913-022-07553-x. PubMed PMID: 35093049; PubMed Central PMCID: PMCPMC8799453.

26. Quispe-Cañari JF, Fidel-Rosales E, Manrique D, Mascaró-Zan J, Huamán-Castillón KM, Chamorro-Espinoza SE, et al. Self-medication practices during the COVID-19 pandemic among the adult population in Peru: A cross-sectional survey. Saudi Pharm J. 2021;29(1):1–11. Epub 20201215. doi: 10.1016/j.jsps.2020.12.001. PubMed PMID: 33519270; PubMed Central PMCID: PMCPMC7832015.

27. Kalam A, Shano S, Khan MA, Islam A, Warren N, Hassan MM, et al. Understanding the social drivers of antibiotic use during COVID-19 in Bangladesh: Implications for reduction of antimicrobial resistance. PLoS One. 2021;16(12):e0261368. Epub 20211214. doi: 10.1371/journal.pone.0261368. PubMed PMID: 34905563; PubMed Central PMCID: PMCPMC8670684.

28. Jirjees F, Ahmed M, Sayyar S, Amini M, Al-Obaidi H, Aldeyab MA. Self-Medication with Antibiotics during COVID-19 in the Eastern Mediterranean Region Countries: A Review. Antibiotics (Basel). 2022;11(6). Epub 20220530. doi: 10.3390/antibiotics11060733. PubMed PMID: 35740140; PubMed Central PMCID: PMCPMC9219972.

29. Shrestha N, Manandhar S, Maharjan N, Twati D, Dongol S, Basnyat B, et al. Perspectives of pharmacy employees on an inappropriate use of antimicrobials in Kathmandu, Nepal. PLoS One. 2023;18(5):e0285287. Epub 20230503. doi: 10.1371/journal.pone.0285287. PubMed PMID: 37134062; PubMed Central PMCID: PMCPMC10156006.

30. Adhikari B, Pokharel S, Raut S, Adhikari J, Thapa S, Paudel K, et al. Why do people purchase antibiotics over-the-counter? A qualitative study with patients, clinicians and dispensers in central, eastern and western Nepal. BMJ Glob Health. 2021;6(5). Epub 2021/05/13. doi: 10.1136/bmjgh-2021-005829. PubMed PMID: 33975888; PubMed Central PMCID: PMCPMC8118002.

31. Rijal KR, Banjara MR, Dhungel B, Kafle S, Gautam K, Ghimire B, et al. Use of antimicrobials and antimicrobial resistance in Nepal: a nationwide survey. Scientific reports. 2021;11(1):11554. Epub 2021/06/04. doi: 10.1038/s41598-021-90812-4. PubMed PMID: 34078956; PubMed Central PMCID: PMCPMC8172831.

32. Kalam MA, Shano S, Afrose S, Uddin MN, Rahman N, Jalal FA, et al. Antibiotics in the Community During the COVID-19 Pandemic: A Qualitative Study to Understand Users’ Perspectives of Antibiotic Seeking and Consumption Behaviors in Bangladesh. Patient Prefer Adherence. 2022;16:217–33. Epub 20220128. doi: 10.2147/ppa.S345646. PubMed PMID: 35115769; PubMed Central PMCID: PMCPMC8806049.

33. Singh DR, Sunuwar DR, Shah SK, Karki K, Sah LK, Adhikari B, et al. Impact of COVID-19 on health services utilization in Province-2 of Nepal: a qualitative study among community members and stakeholders. BMC Health Serv Res. 2021;21(1):174. Epub 20210224. doi: 10.1186/s12913-021-06176-y. PubMed PMID: 33627115; PubMed Central PMCID: PMCPMC7903406.

34. Mertens G, Gerritsen L, Duijndam S, Salemink E, Engelhard IM. Fear of the coronavirus (COVID-19): Predictors in an online study conducted in March 2020. J Anxiety Disord. 2020;74:102258. Epub 20200610. doi: 10.1016/j.janxdis.2020.102258. PubMed PMID: 32569905; PubMed Central PMCID: PMCPMC7286280.

35. Nortey RA, Kretchy IA, Koduah A, Buabeng KO. Biopsychosocial analysis of antibiotic use for the prevention or management of COVID-19 infections: A scoping review. Res Social Adm Pharm. 2023;19(4):573–81. Epub 20221202. doi: 10.1016/j.sapharm.2022.11.011. PubMed PMID: 36496334; PubMed Central PMCID: PMCPMC9715464.

36. Shrestha AB, Aryal M, Magar JR, Shrestha S, Hossainy L, Rimti FH. The scenario of self-medication practices during the covid-19 pandemic; a systematic review. Annals of Medicine and Surgery. 2022:104482.

37. Yuan K, Huang X-L, Yan W, Zhang Y-X, Gong Y-M, Su S-Z, et al. A systematic review and meta-analysis on the prevalence of stigma in infectious diseases, including COVID-19: a call to action. Molecular psychiatry. 2022;27(1):19–33.

38. Rewerska-Juśko M, Rejdak K, editors. Social stigma of patients suffering from COVID-19: challenges for health care system. Healthcare; 2022: MDPI.

39. Charan J, Bhardwaj P, Dutta S, Kaur R, Bist SK, Detha MD, et al. Use of Complementary and Alternative Medicine (CAM) and Home Remedies by COVID-19 Patients: A Telephonic Survey. Indian J Clin Biochem. 2021;36(1):108–11. Epub 20201031. doi: 10.1007/s12291-020-00931-4. PubMed PMID: 33162692; PubMed Central PMCID: PMCPMC7602770.

40. Orisakwe OE, Orish CN, Nwanaforo EO. Coronavirus disease (COVID-19) and Africa: acclaimed home remedies. Scientific African. 2020;10:e00620.

41. Malapela RG, Thupayagale-Tshweneagae G, Baratedi WM. Use of home remedies for the treatment and prevention of coronavirus disease: An integrative review. Health Sci Rep. 2023;6(1):e900. Epub 20221212. doi: 10.1002/hsr2.900. PubMed PMID: 36519078; PubMed Central PMCID: PMCPMC9742825.

42. Hedima EW, Adeyemi MS, Ikunaiye NY. Community Pharmacists: On the frontline of health service against COVID-19 in LMICs. Res Social Adm Pharm. 2021;17(1):1964–6. Epub 20200417. doi: 10.1016/j.sapharm.2020.04.013. PubMed PMID: 32317154; PubMed Central PMCID: PMCPMC7162785.

43. Akour A, Elayeh E, Tubeileh R, Hammad A, Ya’Acoub R, Al-Tammemi AB. Role of community pharmacists in medication management during COVID-19 lockdown. Pathog Glob Health. 2021;115(3):168–77. Epub 20210211. doi: 10.1080/20477724.2021.1884806. PubMed PMID: 33573528; PubMed Central PMCID: PMCPMC8079017.

44. Dutta S, Kaur RJ, Bhardwaj P, Ambwani S, Godman B, Jha PA, et al. Demand of COVID-19 medicines without prescription among community pharmacies in Jodhpur, India: Findings and implications. Journal of family medicine and primary care. 2022;11(2):503–11. Epub 20220216. doi: 10.4103/jfmpc.jfmpc_1250_21. PubMed PMID: 35360769; PubMed Central PMCID: PMCPMC8963618.

45. Elayeh E, Akour A, Haddadin RN. Prevalence and predictors of self-medication drugs to prevent or treat COVID-19: Experience from a Middle Eastern country. Int J Clin Pract. 2021;75(11):e14860. Epub 20210919. doi: 10.1111/ijcp.14860. PubMed PMID: 34516713; PubMed Central PMCID: PMCPMC8646359.

46. Khadka D, Dhamala MK, Li F, Aryal PC, Magar PR, Bhatta S, et al. The use of medicinal plants to prevent COVID-19 in Nepal. J Ethnobiol Ethnomed. 2021;17(1):26. Epub 20210408. doi: 10.1186/s13002-021-00449-w. PubMed PMID: 33832492; PubMed Central PMCID: PMCPMC8027983.

47. Mahato P, Adhikari B, Marahatta SB, Bhusal S, Kunwar K, Yadav RK, et al. Perceptions around COVID-19 and vaccine hesitancy: A qualitative study in Kaski district, Western Nepal. PLOS Glob Public Health. 2023;3(2):e0000564. Epub 20230217. doi: 10.1371/journal.pgph.0000564. PubMed PMID: 36962942; PubMed Central PMCID: PMCPMC10022296.

48. Malik M, Tahir MJ, Jabbar R, Ahmed A, Hussain R. Self-medication during Covid-19 pandemic: challenges and opportunities. Drugs Ther Perspect. 2020;36(12):565–7. Epub 20201003. doi: 10.1007/s40267-020-00785-z. PubMed PMID: 33041621; PubMed Central PMCID: PMCPMC7532737.

49. Ukuhor HO. The interrelationships between antimicrobial resistance, COVID-19, past, and future pandemics. Journal of Infection and Public Health. 2021;14(1):53-60.

50. Kiragga AN, Najjemba L, Galiwango R, Banturaki G, Munyiwra G, Iwumbwe I, et al. Community purchases of antimicrobials during the COVID-19 pandemic in Uganda: An increased risk for antimicrobial resistance. PLOS Glob Public Health. 2023;3(2):e0001579. Epub 20230223. doi: 10.1371/journal.pgph.0001579. PubMed PMID: 36963050; PubMed Central PMCID: PMCPMC10021632.

51. Li J, Zhou P, Wang J, Li H, Xu H, Meng Y, et al. Worldwide dispensing of non-prescription antibiotics in community pharmacies and associated factors: a mixed-methods systematic review. Lancet Infect Dis. 2023. Epub 20230424. doi: 10.1016/s1473-3099(23)00130-5. PubMed PMID: 37105212.

52. Colaneri M, Valsecchi P, Vecchia M, Di Filippo A, Zuccaro V, Seminari E, et al. What prompts clinicians to start antibiotic treatment in COVID-19 patients? An Italian web survey helps us to understand where the doubts lie. J Glob Antimicrob Resist. 2021;26:74–6. Epub 20210609. doi: 10.1016/j.jgar.2021.05.014. PubMed PMID: 34118478; PubMed Central PMCID: PMCPMC8187738.

53. Ghiga I, Pitchforth E, Stålsby Lundborg C, Machowska A. Family doctors’ roles and perceptions on antibiotic consumption and antibiotic resistance in Romania: a qualitative study. BMC Primary Care. 2023;24(1):1–15.

54. Vilovic T, Bozic J, Vilovic M, Rusic D, Zuzic Furlan S, Rada M, et al. Family physicians’ standpoint and mental health assessment in the light of COVID-19 pandemic—a nationwide survey study. International Journal of Environmental Research and Public Health. 2021;18(4):2093.

55. Marcolino MS, Ziegelmann PK, Souza-Silva MV, Nascimento IJBd, Oliveira LM, Monteiro LS, et al. Clinical characteristics and outcomes of patients hospitalized with COVID-19 in Brazil: Results from the Brazilian COVID-19 registry. International Journal of infectious diseases. 2021;107:300–10.

56. Abelenda-Alonso G, Padullés A, Rombauts A, Gudiol C, Pujol M, Alvarez-Pouso C, et al. Antibiotic prescription during the COVID-19 pandemic: A biphasic pattern. Infect Control Hosp Epidemiol. 2020;41(11):1371–2. Epub 20200730. doi: 10.1017/ice.2020.381. PubMed PMID: 32729437; PubMed Central PMCID: PMCPMC7426604.

57. Adhikari B, Yeong Cheah P, von Seidlein L. Trust is the common denominator for COVID-19 vaccine acceptance: A literature review. Vaccine X. 2022;12:100213. Epub 2022/10/12. doi: 10.1016/j.jvacx.2022.100213. PubMed PMID: 36217424; PubMed Central PMCID: PMCPMC9536059.

