## Supplementary File 2 for "Use of antimicrobials during the COVID-19 pandemic: a qualitative study among stakeholders in Nepal" (from another version of this preprint: version 1, 2023-06-29)

### 1. APPENDIX I: Interview guide for dispensers

| SSI number: | Location: |
|---|---|

- Use this interview guide for interviews with dispensers (drug store staff/pharmacists/dispensers)
- Take notes during or after the interview for each topic – just the main points (in English)
- Include description of general ambience, non-verbal reactions and the rough degree of their emotions on particular topics
- You can change the questions but ensure that each issue is covered.
- Avoid using closed questions and probe frequently.
- **Obtain demographic data in the sheet below.**

| **SSI Note Takers Sheet** | | | | | |
|---|---|---|---|---|---|
| **Note Takers initials:** | | | | | |
| **Date: DD-MM-YYY** | | | | | |
| **Dispensary location (town/settlement/neighbourhood):** | | | | | |
| **Socio-demographics** | | | | | |
| **Interviewer ID** | **Age** | **Sex** | **Education (years)** | **Remarks** | **Respondent ID** |

1) Explain the study and ask for consent (memorise the verbal consent script as best you can), then record consent when you turn on microphone

*e.g "Thank you for seeing me today…are you happy to take part in this study?*

**Turn the recorder on**

*Ok, so I have turned the microphone on. I just wanted to ask again, are you happy to take part in this study by speaking with me today?*

2) Explore the themes below:

| **OTC: (Definition: Buying "over the counter medicine" means purchasing medicine from a local dispenser who is not the health worker/physician or buying medicine without a prescription)** | |
|---|---|
| **Topic** | **Suggested questions/probes** |
| **Tell me about this dispensary…** | • How many customers do you see in a day?<br>• Is the place yours or do you work for someone?<br>• How long have you been working here?<br>• How long has the shop been open?<br>• Have you seen any changes in this time? What are they? |
| **Main illnesses treated** | • What complaints do your customers suffer?<br>• Has this changed since the start of the pandemic?<br>• What type of medicines do your customers request? |
| **Demand for medicines** | • In general, what are the medication patient spontaneously demand?<br>• What is your general reaction to them?<br>• Do different types of people ask for different medicines?<br>• Do they directly ask for a medicine or is there a more subtle way of asking…e.g. telling a story about friend who was sick and received a certain medicine?<br>• Do you ever ask advice from anyone if you receive these requests?<br>• Has this changed since the start of the pandemic? In what ways? |

| | |
|---|---|
| **Treating respiratory infections** | • When a patient visits you with a cough, can you tell us what do you do?<br>• Do you do lab examination?<br>• Do you consult with someone else?<br>• Do you prescribe antibiotics?<br>• On what basis? when do you call for the follow up?) do patients with fever demand antibiotics?/if so, what do you do?<br>• What about common cold/flu, how do you treat them? Do these patients with flu demand for anitbiotics? If so, what do you do? |
| **Purchasing antibiotics** | • Do you dispense antibiotics?<br>• How often do you dispense antibiotics?<br>• What types of antibiotics?<br>• Do you sell them as individual tablets or as a whole course?<br>• How much do they cost?<br>• Which antibiotic do you sell the most?<br>• What complaints are they sold for?<br>• Do you ever refuse to sell antibiotics?<br>• If you don’t have them in stock can you get them from somewhere else?<br>• Do you ever refer customers to other providers? |
| **Treating COVID-19** | • What do you do if some visits who you suspect might have COVID-19?<br>• Can you explain what kind of signs and symptoms these patients came with?<br>• Had any of these people have a positive test result with them? Did you refer them for a test? |

| | |
|---|---|
| | • Have you ever dispensed medicines to some who you thought might have COVID?<br>• How many times in the last month?<br>• What do you dispense?<br>• Did your treatment involve antibiotics? If yes, what antibiotics did you offer them? Why do you choose those?<br>• Can you tell whether these antibiotics work against COVID-19? If so, can you explain us how they work or how they do not work?<br>• Do you give advice?<br>• Do you think there was a rise in patients attending your clinic due to the pandemic? |
| **Potential consequences of OTC dispensing for COVID** | • Did these antibiotics work for or against the COVID-19 cases?<br>• Do you think COVID-19 pandemic may have increased the sale of antibiotics?<br>• Do you think COVID-19 may have increased the antibiotic pressure or antibiotic resistance? |
| **CLOSING**<br><br>Summarise the key points from the interview | Do you have any questions or concerns you'd like to raise? Thank you. |

### 2. APPENDIX II: Interview guide for patients who were diagnosed with COVID-19

| **SSI number:** | **Location:** |
|---|---|

- Use this interview guide for interviews with patients or their peers, and relatives
- Take notes during or after the interview for each topic – just the main points (in English)
- Include description of general ambience, non-verbal reactions, and the rough degree of their emotions on particular topics
- You can change the questions but ensure that each issue is covered.
- Avoid using closed questions and probe frequently.
- **Obtain demographic data in the sheet below.**

| **SSI Note Takers Sheet** | | | | | |
|---|---|---|---|---|---|
| **Note Takers initials:** | | | | | |
| **Date: DD-MM-YYY** | | | | | |
| **Address of the location (town/settlement/neighbourhood):** | | | | | |
| **Socio-demographics** | | | | | |
| **Interviewer ID** | **Age** | **Sex** | **Education (years)** | **Remarks** | **Respondent ID** |

1) Explain the study and ask for consent (memorise the verbal consent script as best you can), then record consent when you turn on microphone

*e.g "Thank you for seeing me today…are you happy to take part in this study?*

**Turn the recorder on**

*Ok, so I have turned the microphone on. I just wanted to ask again, are you happy to take part in this study by speaking with me today?*

2) Explore the themes below:

| **OTC: (Definition: Buying "over the counter medicine" means purchasing medicine from a local dispenser who is not the health worker/physician or buying medicine without a prescription)** | |
|---|---|
| **Topic** | **Suggested questions/probes** |
| **Tell me about your general condition** | • How are you today?<br>• How has been the last few weeks in terms of your health? |
| **Febrile illnesses OTC** | • Can you share me your experience if you have febrile illnesses recently?<br>• What did you do when you had fever?<br>• Did you seek medications from drug store?<br>• Do you know what medications you got for your fever? |
| **Demand for medicines** | • In general, what kind of medicine do you buy from the drug shop?<br>• How do you decide what medication to ask in the drug shop?<br>• Do you buy medicine for yourself or for your family members?<br>• Do you ever ask your relatives/neighbours for medicines?<br>• What about during the pandemic? Did you seek anyone's help to buy medicines? |

| | |
|---|---|
| **Treating respiratory infections** | • When you have a cough, can you tell us what do you do?<br>• Do you do lab test for your condition?<br>• Do you self-medicate with any medicines or antibiotics?<br>• What about common cold/flu, what do you do to cure it? Do you take medicines when you have flu? If so, what do you do? |
| **Purchasing antibiotics** | • Do you buy specific medicines such as antibiotics?<br>• How often do you buy antibiotics?<br>• What types of antibiotics do you buy?<br>• Do you generally buy medicine for full course or for only few days?<br>• How much do they cost?<br>• Which antibiotic do you buy the most?<br>• What conditions do you often buy the antibiotics for?<br>• Do you ever think not to buy antibiotics? Why? |
| **COVID-19 and OTC**<br>**(Perceived/projected attitude when a person gets COVID-19)** | • Have you seen/noticed any of your relatives/peers/friends who suffered from COVID-19?<br>• Based on your observation/experience, can you explain us how the symptoms and signs of COVID-19 begin and progress?<br>• Do you think all patients with COVID-19 seem to suspect or know their disease?<br>• Do these patients want to know their condition by lab confirmation? (if not why?)<br>• What do these patients with symptoms and signs of COVID-19 do to mitigate the severity?<br>• Do you think these patients buy medication over the counter?<br>• What kind of medications do they buy from the drug shops? |

| | |
|---|---|
| | • Do you think they receive antibiotics for their symptoms? |
| **COVID-19 and OTC (lived experience)** | • Before we talk about COVID-19 diagnosis, can you explain me what were the initial symptoms and signs and how it progressed?<br>• What did you do to mitigate the initial signs/symptoms?<br>• Before the test, did you suspect that it could be COVID-19?<br>• So, how did you decide that you would want to do the test?<br>• Initially and for the mild symptoms, what medications did you use? Where did you get them from? Can you explain, how did you get them?<br>• Do you know what medicine the drug shop provided to you? were they antibiotics? |
| **Potential consequences of OTC dispensing for COVID** | • Do you think receiving antibiotics from over the counter without diagnosis of COVID-19 helpful?<br>• What are the benefits and disadvantages of OTC medications for patients who are suspected/diagnosed with COVID-19?<br>• Do you think antibiotics work for or against the COVID-19 cases?<br>• Do you think COVID-19 pandemic may have increased the demands for antibiotics?<br>• Do you think COVID-19 may have increased the antibiotic pressure or antibiotic resistance? |
| **CLOSING**<br>Summarise the key points from the interview | Do you have any questions or concerns you'd like to raise? Thank you. |

### 3. APPENDIX III: Interview guide for health workers

| SSI number: | Location: |
|---|---|

- Use this interview guide for interviews with health workers (clinicians who are responsible for the treatment of COVID-19)
- Take notes during or after the interview for each topic – just the main points (in English)
- Include description of general ambience, non-verbal reactions and the rough degree of their emotions on particular topics
- You can change the questions but ensure that each issue is covered.
- Avoid using closed questions and probe frequently.
- **Obtain demographic data in the sheet below.**

| **SSI Note Takers Sheet** | | | | | |
|---|---|---|---|---|---|
| **Note Takers initials:** | | | | | |
| **Date: DD-MM-YYY** | | | | | |
| **Dispensary location (town/settlement/neighbourhood):** | | | | | |
| **Socio-demographics** | | | | | |
| **Interviewer ID** | **Age** | **Sex** | **Education (years)** | **Remarks** | **Respondent ID** |

1) Explain the study and ask for consent (memorise the verbal consent script as best you can), then record consent when you turn on microphone

*e.g "Thank you for seeing me today…are you happy to take part in this study?*

**Turn the recorder on**

*Ok, so I have turned the microphone on. I just wanted to ask again, are you happy to take part in this study by speaking with me today?*

2) Explore the themes below:

| **OTC: (Definition: Buying "over the counter medicine" means purchasing medicine from a local dispenser who is not the health worker/physician or buying medicine without a prescription)** | |
|---|---|
| **Topic** | **Suggested questions/probes** |
| **Tell me about your setting** | • How many patients do you see in a day?<br>• How long have you been working here?<br>• How long has the clinics/hospital been open?<br>• Have you seen any changes in this time? What are they? |
| **Main illnesses treated** | • What complaints do your patients suffer?<br>• Has this changed since the start of the pandemic?<br>• What type of medicines do your patients request? |
| **Demand for medicines** | • In general, what are the medication patient spontaneously demand?<br>• What is your general reaction to them?<br>• Do different types of patients ask for different medicines?<br>• Has this changed since the start of the pandemic? In what way? |

| | |
|---|---|
| **Treating respiratory infections** | • When a patient visits you with a cough, can you tell us what do you do?<br>• Do you do lab examination?<br>• Do you consult with someone else?<br>• Do you prescribe antibiotics?<br>• On what basis? when do you call for the follow up?) do patients with fever demand antibiotics?/if so, what do you do?<br>• What about common cold/flu, how do you treat them? Do these patients with flu demand for anitbiotics? If so, what do you do? |
| **Purchasing antibiotics** | • Do you think patients buy antibiotics over the counter?<br>• What types of antibiotics?<br>• Do dispensers sell them as individual tablets or as a whole course?<br>• How much do they cost?<br>• Which antibiotic do you think they sell the most?<br>• What complaints are they sold for?<br>• Do you think they ever refuse to sell antibiotics? |
| **Treating COVID-19** | • What do you do if someone who you suspect might have COVID-19?<br>• Can you explain what kind of signs and symptoms these patients came with?<br>• Had any of these people have a positive test result with them? Did you refer them for a test?<br>• How many times in the last month?<br>• Did your treatment involve antibiotics? If yes, what antibiotics did you offer them? Why do you choose those? |

| | |
|---|---|
| | • Can you tell whether these antibiotics work against COVID-19? If so, can you explain us how they work or how they do not work?<br>• Do you give advice?<br>• Do you think there was a rise in patients attending your clinic due to the pandemic? |
| **Potential consequences of OTC dispensing for COVID** | • Did these antibiotics work for or against the COVID-19 cases?<br>• Do you think COVID-19 pandemic may have increased the sale of antibiotics?<br>• Do you think COVID-19 may have increased the antibiotic pressure or antibiotic resistance? |
| **CLOSING**<br>Summarise the key points from the interview | Do you have any questions or concerns you'd like to raise? Thank you. |
